# Measuring Adherence to Multiple Medications Using Guideline-Directed Medical Therapy as a Model

**DOI:** 10.64898/2025.12.05.25341740

**Authors:** Eli Reynolds, Xiyue Li, Amrita Mukhopadhyay, Samrachana Adhikari, Carine E. Hamo, Adam Berman, Morgan E. Grams, Saul Blecker

## Abstract

**Background:** There is no gold standard for measuring adherence to a complex medication regimen. Heart failure is a chronic disease state that requires multiple medications for optimal control, known as guideline-directed-medical therapy (GDMT), and can be used a model to explore approaches to assessing multi-medication adherence. We aimed to compare seven proportion of days covered (PDC) measures for assessing adherence to multiple GDMT medications and to evaluate their association with clinical outcomes.

**Methods:** We conducted a large, single center, retrospective cohort study of 34,603 patients with heart failure who filled a GDMT prescription between April 2021 and July 2022. The primary outcomes were the seven PDC-based adherence measures derived from electronic health record and pharmacy data. PDC was defined as the proportion of days in which patients had possession of GDMT medications. Our secondary outcome was the combined clinical outcome of ED visits, hospitalizations, and death.

**Results:** The seven measures provided a wide range of measured PDC adherence. Using the strictest measure, ‘each’ (>= 80% of days with each drug available) 54% of patients were considered adherent, compared with 73% measured adherence for the least restrictive measure, ‘at least one’. Variability increased with increasing number of medications across all non-average based measures. For example, using the ‘all’ measure (a more restrictive PDC measure) adherence ranged from 0.53 to 0.41 with increasing number of GDMT prescriptions. Higher PDC for each of the seven measures was associated with increased number of ED, visits, hospitalizations, and death. There was no association with the combined outcome in patients with heart failure with reduced ejection fraction.

**Conclusions and Relevance:** There was a wide variability in adherence measures for assessing adherence to GDMT depending on the measure used. This variability has significant implications for the policy, clinical, and intervention context to which the measure is applied.

## INTRODUCTION

Medication adherence is associated with important patient health outcomes, including hospitalizations and death (1,2). Measurements of medication adherence have been used for quality improvement and reimbursement programs, clinical and population health interventions and research (3,4). Confidence in assessment of adherence is fundamental as a starting point across these domains. The proportion of days covered (PDC) is one of the most utilized measures of medication adherence at the population level and has been endorsed as a quality metric by the Pharmacy Quality Alliance (PQA) (5–8). Traditionally, PDC employs insurance-based pharmacy refill claims to estimate the proportion of days a person has access to a given medication over a specified period of interest (6). When compared to other measures such as pill count or Medication Event Monitoring Systems (MEMS), PDC is a more feasible approach to measuring population-based adherence (5). PDC typically has been utilized to assess adherence for one or two medication classes. Only a limited number of studies have applied PDC to multiple medications or medication classes, despite the fact that many chronic diseases have recommended multi-drug regimens (9–11). Therefore, it is currently unclear how to calculate PDC if more than two drugs are involved, and various approaches could be considered.

Heart failure is a chronic condition associated with a high morbidity and mortality that requires multiple medications to effectively manage. Specifically, adherence to guideline-directed-medical-therapy (GDMT) is core to the therapeutic approach (12,13). The four GDMT classes for HFrEF include renin-angiotensin system inhibitors (RAASi), evidence-based beta-blockers, mineralocorticoid inhibitors (MRA), and sodium-glucose cotransporter 2 inhibitors (SGLT2i).

These heart failure medications have shown reduction in mortality and hospitalizations in randomized control trials (12,13). Despite the importance of adhering to these medications, the optimal approach to calculating PDC for these four classes of medications has not been well described.

The purpose of this study was to describe and compare seven different approaches to assessing adherence for multiple medications, using heart failure as a model. We calculated PDC for each of these approaches. Our secondary objective was to assess the association of each approach with important clinical outcomes in heart failure.

## METHODS

We conducted a retrospective cohort study of patients with heart failure at New York University (NYU) Langone Health, a large, multi-site health system that includes six acute care hospitals and over 300 ambulatory clinics in greater New York City and Florida. Our primary data source was the EHR at NYU, Epic (Epic Systems, Verona, WI). The EHR contains patient demographic information, inpatient and outpatient encounter information, basic clinical and laboratory variables, and medication orders. EHR data was linked to pharmacy and pharmacy benefits manager fill data through SureScripts (Surescripts, Arlington, VA) to obtain medication fill information (14). This study was approved by the NYU Institutional Review Board with a waiver of informed consent.

The study included adults aged > 18 years old with a diagnosis of heart failure [as defined by the International Classification of Diseases, 10^th^ Revision codes (15)] or those with an ejection fraction less than or equal to 40% and had at least one clinical encounter between April 1, 2021, and July 31, 2022. Participants were required to have at least one active prescription out of the four GDMT classes: RAASi, beta blocker, non-steroidal MRA or SGLT-2i. The index date for each participant was defined as the earliest date of prescription within the study period with the highest number of GDMT medication classes prescribed (9,10).

Our primary measure of interest was medication adherence, assessed as PDC. PDC was defined as the ratio of the number of days that a medication was available, based on pharmacy fills, over the number of days with an active prescription (5). We calculated PDC over a 180-day period starting with the index date, which was defined as the first date within the study period with the highest number of active GDMT prescriptions. EHR medication order data determined whether a particular medication was active (16). Lapses in active prescription status as recorded by the EHR were not included in the denominator. Additionally, we excluded inpatient hospitalizations from the denominator. Medications within a specific GDMT class were considered interchangeable.

We used seven different approaches to calculate PDC based on prior work (*Table 1*) (17). We used three continuous measures of PDC for multiple medications: ‘average,’ ‘maximum,’ and ‘minimum.’ ‘Average’ was defined as the sum of each medication’s PDC over number of GDMT medication classes. ‘Maximum’ was defined as days with at least one GDMT medication available divided by active prescription days in the time interval. Similarly, ‘minimum’ was defined as the proportion of days with all medications available. We employed four dichotomous measures of adherence, in which a patient was considered adherent if the PDC was greater than or equal to 80%, as is standard (17). In the ‘at least one’ measure a patient was defined as adherent if 80% of days were covered by one or more GDMT medication. In contrast, the ‘all’ definition was defined as over 80% of days with all prescribed GDMT medications. The ‘each’ definition, the strictest measure, of adherence was based on whether each GDMT medication was available for at least 80% of active prescription days. Finally, in the ‘average-each’ definition a patient was adherent if the average of the individual medication adherence rates of each prescribed class was at least 80%.

**Table 1.** Seven distinct PDC-based definitions of adherence to multiple medications using GDMT as an example.

| Measure | Definition | Formula for PDC calculation |
| --- | --- | --- |
| *Continuous* |  |  |
| Minimum | Proportion of days covered by all GDMT medications | $\left[ \frac{\text{Dayswithallmedicationsavailable}}{\text{DayswithActiveRx}} \right]$ |
| Maximum | Proportion of days covered by at least one medication | $\frac{\text{Dayswith} \geq 1 \text{ medicationavailable}}{\text{DayswithActiveRx}}$ |
| Average | Average of adherence rates of individual medications | $\left[ \frac{\text{DayswithRAAI}}{\text{DayswithActiveRx}} + \frac{\text{DayswithSGLT2i}}{\text{DayswithActiveRx}} + \frac{\text{DayswithMRA}}{\text{DayswithActiveRx}} + \frac{\text{Dayswith}\beta \text{ blocker}}{\text{DayswithActiveRx}} \right] / \# \text{ofActiveRxclasses}$ |
| *Dichotomous* |  |  |
| At least one | Adherent if $\geq 80\%$ of days covered by at least one medication | Adherent if $\left[ \frac{\text{Dayswith} \geq 1 \text{ medicationavailable}}{\text{DayswithActiveRx}} \right] \geq 80\%$ |
| All | Adherent if $\geq 80\%$ of days covered by at least one medication | Adherent if $\left[ \frac{\text{Dayswithallmedicationsavailable}}{\text{DayswithActiveRx}} \right] \geq 80\%$ |
| Each | Adherent if each of multiple medications covers $\geq 80\%$ of days | Adherent if $\left[ \begin{aligned} &\frac{\text{DayswithRAAI}}{\text{DayswithActiveRx}} \geq 80\% \text{ and } \frac{\text{DayswithSGLT2i}}{\text{DayswithActiveRx}} \geq 80\% \text{ and } \frac{\text{DayswithMRA}}{\text{DayswithActiveRx}} \geq 80\% \text{ and } \frac{\text{Dayswith}\beta \text{ blocker}}{\text{DayswithActiveRx}} \geq 80\% \end{aligned} \right]$ |
| Average-Each | Adherent if average of individual medication | $\left[ \frac{\text{DayswithRAAI}}{\text{DayswithActiveRx}} + \frac{\text{DayswithSGLT2i}}{\text{DayswithActiveRx}} + \frac{\text{DayswithMRA}}{\text{DayswithActiveRx}} + \frac{\text{Dayswith}\beta \text{ blocker}}{\text{DayswithActiveRx}} \right] / \# \text{ofActiveRxclasses} \geq 80\%$ |
| | adherence rates ≥ 80% | Average of adherence rates to individual medications,<br>Adherent if ≥ 80% of days |

RAAI = Renin Angiotensin Aldosterone Inhibitor SGLT2i = Sodium-glucose cotransporter 2 inhibitor MRA = Mineralocorticoid receptor antagonist $\beta$ blocker = beta blocker Rx = prescription

### Clinical Outcomes

For our secondary objective, we assessed the association of each our seven approaches to a composite clinical outcome. The exposures for this analysis were the calculated PDC for each patient. The primary outcome was the combination of all-cause hospitalization, ED visits, and death assessed starting at time of PDC measurement (which reflected the prior six months of medication fills) through the study end date on 30 April 2023. Participants were excluded if death occurred during the six-month PDC period.

For this analysis, we included patients with two or more prescription classes, given our interest in the performance of the measures for multiple medications. Covariates including age, sex, race and ethnicity, language, insurance status, community type, Elixhauser comorbidity index, number of GDMT medications, most recent ejection fraction for a patient, history of reduced ejection fraction, total hospitalization days, and number of outpatient visits in the past year (*Table 1*).

### Statistical Analysis

Baseline descriptive characteristics were tabulated for the study sample and expressed as appropriate measures of central tendency and frequencies. Our primary measures of PDC for each of the seven definitions were presented for the entire cohort and also by the number of GDMT medication classes. Our measure of PDC was expressed as a mean (SD) for the continuous definitions, while frequency was used for the binary definitions.

In our secondary analysis, the continuous measures of PDC were analyzed as tertiles. If tertiles were unable to be created due to the median measure of PDC of one, the measure was analyzed as above and below the median. For the assessment of the association of PDC approaches and the composite clinical outcomes, we conducted an unadjusted time-to-event Cox proportional hazard analysis employing a left-truncation model to estimate hazard ratios (HR) and 95% confidence intervals (CI). We used the seven approaches of adherence as the independent variables and composite outcome of all-cause hospitalizations, ED visits, and death as the dependent variable.

To account for potential confounders for the primary combined outcome, adjustments were made using the baseline covariates. Furthermore, an unadjusted and adjusted subgroup analysis was performed for patients with a reduced ejection fraction. Multivariate regression models were employed to account for any confounding or effect-modifying variables. Two-sided hypothesis testing at an alpha level of 0.05 was used for all significance testing. We used R version 4.2.2 (R Foundation for Statistical Computing) as the statistical software of choice to perform our analysis.

## RESULTS

### Patient Characteristics

The study included 34,603 patients with heart failure. The demographic characteristics of our population are shown in *Table 2*. Roughly half were male (56.2%), and the average age was 73.1 (standard deviation (SD) 13.4) years. The self-reported race/ethnicity was 64.5% white, 13.1% Black, 10.3% Hispanic, and 4.0% Asian. English was the primary language for 84.1% of the participants. Regarding insurance status, 71% of our cohort was insured by Medicare, 20% by commercial insurance, and then finally 8% by Medicaid. Our cohort was almost entirely metropolitan (99%). Our population had a mean (SD) Elixhauser comorbidity index of 12.9 (7.94).

**Table 2.**
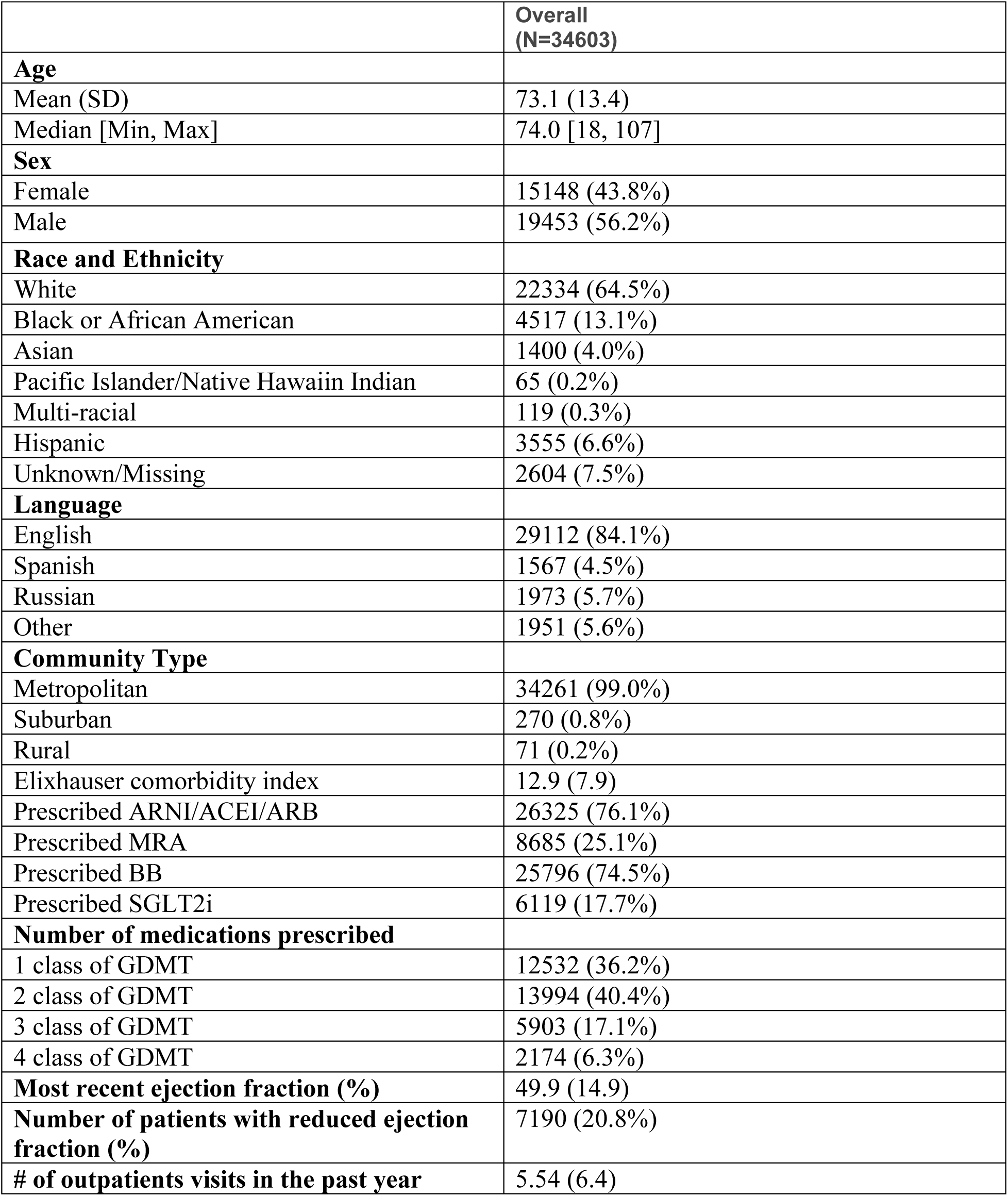
Baseline demographic characteristics of study cohort.

### Adherence According to Each Definition

The most commonly prescribed GDMT was RAAS inhibitors (76.1%), followed by beta blockers (74.5%), mineralocorticoid antagonists (25.1%), and SGLT2 inhibitors (17.7%). Among patients, 12,532 (36.2%) were prescribed 1 class of GDMT, 13,994 (40.4%) were prescribed 2 classes, 5903 (17.1%) were prescribed 3 classes, and 2174 (6.3%) were prescribed 4 classes.

The calculated PDC using the seven approaches to multiple medications are shown in (*Table 3)*. The PDC for continuous definitions, ranged from a mean (standard deviation) of 0.68 (0.37) for the ‘minimum’ definition to 0.81 (0.32) for the ‘maximum’ definition; the mean PDC using the ‘average’ definition was 0.74 (0.32). Within the dichotomous definition group, the ‘all’ definition had an adherence rate of 54.3%, the ‘at least one’ definition had a rate of 73.1%, the ‘each’ definition had a rate of 53.5%, and the ‘average-each’ definition had a rate of 59.9%.

**Table 3.** Medication adherence as measured by proportion of days covered (PDC) and definitions for approach to multiple medication classes. Results are presented for both the overall cohort and subgroups of patients based on number of GDMT medication classes.

|  | **Overall Cohort**<br>**(N=34603)** | **1 GDMT Class**<br>**(N=12532)** | **2 GDMT Classes**<br>**(N=13994)** | **3 GDMT Classes**<br>**(N=5903)** | **4 GDMT Classes**<br>**(N=2174)** |
| --- | --- | --- | --- | --- | --- |
| *Continuous Mean (SD)* |  |  |  |  |  |
| **Minimum** | 0.676<br>(0.365) | 0.728<br>(0.358) | 0.670<br>(0.364) | 0.607<br>(0.367) | 0.596<br>(0.358) |
| **Maximum** | 0.808<br>(0.320) | 0.728<br>(0.358) | 0.836<br>(0.300) | 0.871<br>(0.273) | 0.922<br>(0.218) |
| **Average** | 0.739<br>(0.323) | 0.728<br>(0.358) | 0.744<br>(0.311) | 0.740<br>(0.290) | 0.768<br>(0.254) |
| *Dichotomous, %* |  |  |  |  |  |
| **All** | 18793<br>(54.3%) | 7831<br>(62.5%) | 7471<br>(53.4%) | 2599<br>(44.0%) | 892<br>(41.0%) |
| **At least one** | 25292<br>(73.1%) | 7831<br>(62.5%) | 10746<br>(76.8%) | 4789<br>(81.1%) | 1926<br>(88.6%) |
| **Each** | 18500<br>(53.5%) | 7831<br>(62.5%) | 7371<br>(52.7%) | 2482<br>(42.0%) | 816<br>(37.5%) |
| **Average-Each** | 20716<br>(59.9%) | 7831<br>(62.5%) | 8348<br>(59.7%) | 3285<br>(55.6%) | 1252<br>(57.6%) |

We then calculated PDC for each approach to multiple medication adherence, among subgroups of patients on one, two, three, or four classes of GDMT (*Table 3*). All seven definitions produced the same adherence measurement and for one medication class: 0.73 (0.36) for the continuous measures and 62.5% for the dichotomous measures. Within the continuous definitions, the ‘maximum’ definition produced a higher adherence rate with increasing number of medications, with PDC of 0.84 (0.30) for two medications, 0.87 (0.27) for three medications, and 0.92 (0.22) for four medications. The opposite trend was observed for the ‘minimum’ definition with PDC of 0.67 (0.36), 0.61 (0.38), and 0.60 (0.36) for two, three, and four medications, respectively. Conversely, the ‘average’ category stayed relatively stable across number of medications, ranging from 0.74 (0.29) for two medication classes to 0.77 (0.25) for four medication classes. We observed similar patterns across different dichotomous definitions, with calculated PDC decreasing with increasing numbers of medications using the ‘all’ and ‘each’ (i.e. 62.5% for 1 GDMT class to 41.0% for 4 GDMT classes for the ‘all’ definition). Conversely, PDC increased with increasing number of medications using the ‘at least one’ definition, from 62.5% to 88.6%. PDC calculated using the ‘average-each’ definition was more consistent across numbers of medications (*Table 3*).

### Clinical Outcomes

The outcome analysis included 22,071 (63.8%) patients after removing those with only one medication class. In an unadjusted model, each of the four dichotomous measures of PDC (at least one, all, each, and average-each) were associated with a significantly higher risk of the composite outcome of ED visits, hospitalizations, and death. Specifically, the ‘at least one’, ‘all, ‘each’ and ‘average-each’ had hazard ratios of 1.18 (1.11-1.25), 1.06 (1.02-1.12), 1.08 (1.03-1.14), and 1.07 (1.02-1.12), respectively when comparing the highest tertile of adherence versus the lowest tertile. These associations were similar in a model adjusted for baseline covariates, hospitalization days, number of GDMT medications, and number of outpatient cardiology visits. (*Table 4A*). In an unadjusted model, each of the three continuous measures of PDC (minimum, maximum, and average) were associated with a significantly higher risk of the composite outcome of ED visits, hospitalizations, and death (*Table 4B*).

**Table 4A.** Association of the four dichotomous PDC adherence measures with the combined outcome of ED visit, all-cause hospitalization, and death.

| Measurement | Composite Outcome | p-value | Composite Outcome* | p-value |
| --- | --- | --- | --- | --- |
| At least one | 1.18 (1.11-1.25) | <0.001 | 1.18 (1.11-1.26) | <0.001 |
| All | 1.06 (1.02-1.12) | 0.01 | 1.03 (0.98-1.08) | 0.21 |
| Average | 1.08 (1.03-1.14) | 0.001 | 1.06 (1.01-1.12) | 0.02 |
| Average-Each | 1.07 (1.02-1.12) | 0.01 | 1.05 (1.00-1.11) | 0.03 |

\*Adjustments made for covariates of hospitalization, number of GDMT, number of outpatient cardiology visits, age, sex, race and ethnicity, language, insurance, Elixhauser comorbidity score, most recent ejection fraction, and total hospitalization days during follow up.

**Table 4B.** Association of the three continuous PDC adherence measures with the combined clinical outcome in the entire cohort.

|  | PDC 1<sup>st</sup> tertile | PDC 2<sup>nd</sup> tertile | PDC 3<sup>rd</sup> tertile |
| --- | --- | --- | --- |
| *Unadjusted* |  |  |  |
| Minimum | 1.16 (1.08-1.24) | 1.12 (1.05-1.20) | 1.18 (1.10-1.27) |
| Maximum |  | 1.09 (1.03-1.14) | “” |
| Average | 1.14 (1.06-1.22) | 1.13 (1.11-1.21) | 1.18 (1.11-1.27) |
|  | PDC 1<sup>st</sup> tertile | PDC 2<sup>nd</sup> tertile | PDC tertile |
| *Adjusted\** |  |  |  |
| Minimum | 1.11 (1.04-1.19) | 1.08 (1.00-1.15) | 1.11 (1.03-1.19) |
| Maximum** | “” | 1.07 (1.02-1.13) | “” |
| Average | 1.08 (1.004-1.15) | 1.09 (1.02-1.17) | 1.12 (1.04-1.20) |

\*Adjustments made for covariates of hospitalization, number of GDMT, number of outpatient cardiology visits, age, sex, race and ethnicity, language, insurance, Elixhauser comorbidity score, most recent ejection fraction, and total hospitalization days during follow up.
\*\* The maximum definition was made binary given the median for the definition was 1.

In a subgroup analysis of the cohort of patients with HFrEF (N=7190 (20.8%)), we found no significant association of any of the seven measures of PDC with the combined outcome of ED visits, hospitalizations, and death (*Table 4C*). The adjusted analysis demonstrated similar results to the unadjusted analysis (*Table 4D*).

**Table 4C.**
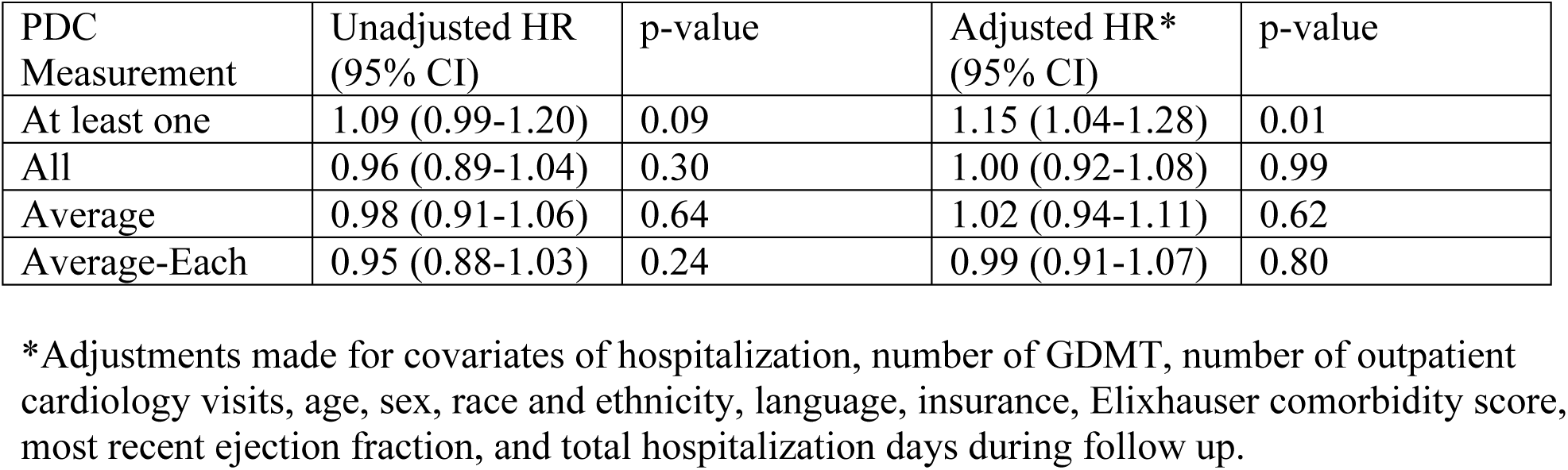
Association of the four dichotomous PDC adherence measures with the combined outcome of ED visits, hospitalizations, and death in patients with heart failure with reduced ejection fraction (N=7190).

| PDC Measurement | Unadjusted HR (95% CI) | p-value | Adjusted HR* (95% CI) | p-value |
| --- | --- | --- | --- | --- |
| At least one | 1.09 (0.99-1.20) | 0.09 | 1.15 (1.04-1.28) | 0.01 |
| All | 0.96 (0.89-1.04) | 0.30 | 1.00 (0.92-1.08) | 0.99 |
| Average | 0.98 (0.91-1.06) | 0.64 | 1.02 (0.94-1.11) | 0.62 |
| Average-Each | 0.95 (0.88-1.03) | 0.24 | 0.99 (0.91-1.07) | 0.80 |

\*Adjustments made for covariates of hospitalization, number of GDMT, number of outpatient cardiology visits, age, sex, race and ethnicity, language, insurance, Elixhauser comorbidity score, most recent ejection fraction, and total hospitalization days during follow up.

**Table 4D.**
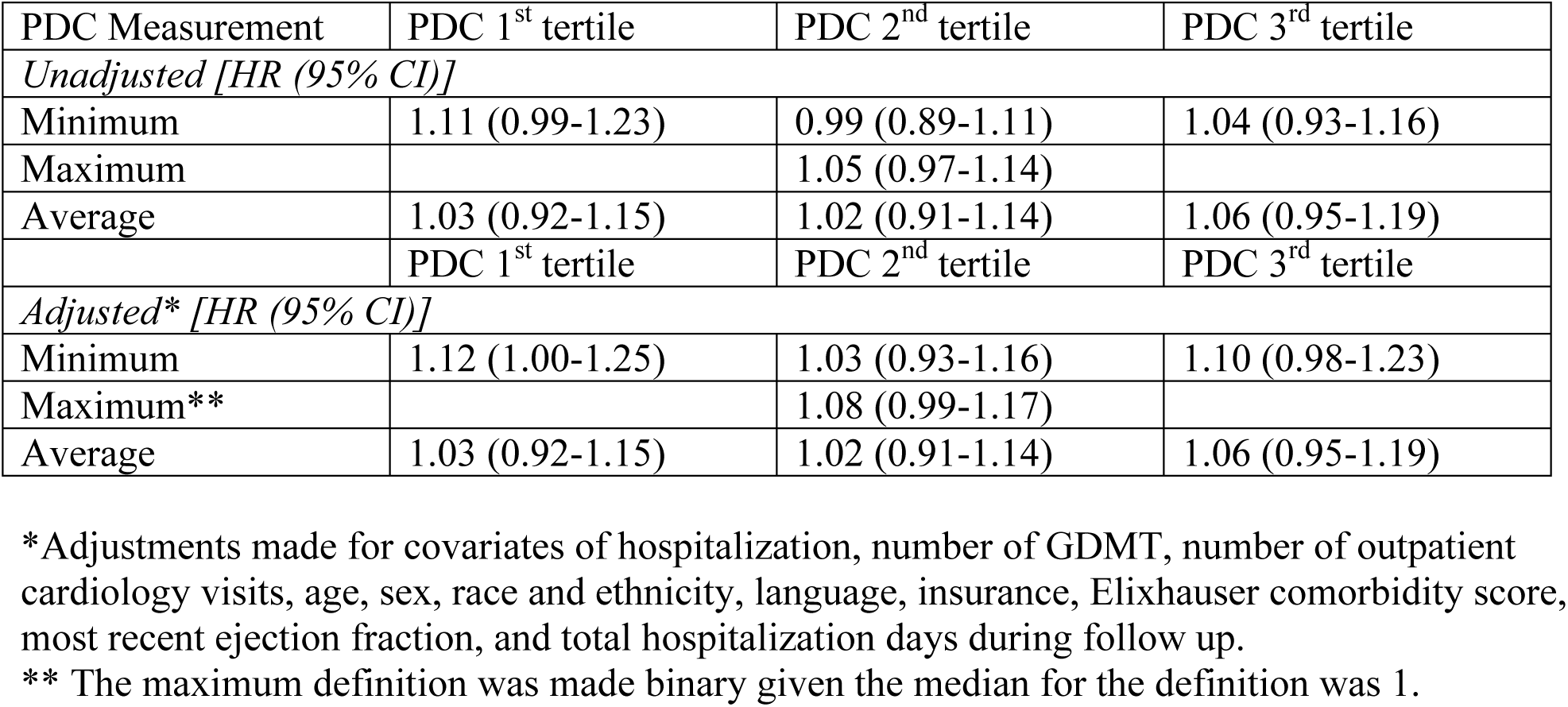
Association of the three continuous PDC adherence measures with the combined clinical outcome of ED visits, all-cause hospitalizations, and death in patients with heart failure with reduced ejection fraction (N=7190).

## DISCUSSION

In this study of over 30,000 patients with heart failure, patients were prescribed on average about two GDMT medications for heart failure. We used seven different approaches to calculate medication adherence for GDMT, as measured by PDC. We found significant variation in PDC depending on the approach. For instance, 54% of patients were considered adherent to therapy using an approach which required adherence to all medications (‘all’ definition) while 73% of patients were considered adherent to therapy if they were adherent to at least one medication (‘at least one’ definition). Surprisingly, we found that increased PDC was associated with increased rates of ED visits, hospitalizations, and mortality regardless of approach used. Of note, this association was not observed among a subgroup of patients with HFrEF. Nonetheless, the outcome association was similar for each of the approaches to calculating PDC using multiple medications.

We found that assessment of adherence based on PDC substantially differed, depending on the measure used to account for multiple medications. When using more restrictive definitions, which required patients to possess all their medication on a given day to be considered adherent, we estimated that patients were adhering to prescribed therapy between half and two-thirds of the time. Conversely, when using less restrictive measures of adherence, which considered a patient to be adherent if they were taking at least one medication on a given day, we estimated adherence in our population to be 10-20% higher. Similar PDC measurement trends were seen in prior adherence work, though these were limited by looking at only two medication classes and our work suggests this range is even wider as more medications are added (9–11).

In addition to variability between definitions, we found that the number of medications led to variability within each definition. Specifically, adding more medications led to even lower estimates of the more restrictive definitions for multiple medications. For instance, the ‘minimum’ approach derived a PDC adherence measure that ranged from 0.73 with one medication to 0.60 with four medications, a finding which may reflect additional challenges for patients to make sure they are taking all medications every day as the number of medications increase. Conversely, less restrictive definitions increased with increasing number of medications. For example, the ‘maximum’ definition of PDC ranged from 0.73 for one medication class to 0.92 for four medication classes, a finding that may reflect increased opportunity for patients to ensure they were taking at least one medication regularly. We observed a smaller range in adherence measurements in the average definitions when compared with the cumulative definitions in both the dichotomous and continuous definitions.

These differences in estimates have important implications for measurement of adherence using PDC with multiple medications. Notably, such measurement has been used for quality improvement efforts and as a quality metric. For example, The Centers for Medicare & Medicaid Services (CMS) uses PDC measurement for quality bonuses for diabetes in its STAR rating program, a system developed by CMS to rate the quality of Medicare Advantage (Part C) and Prescription Drug (Part D) plans (18,19). These quality incentives are projected to be valued around 6 billion annually (20). Currently, CMS uses the ‘at least one’ approach in measuring adherence for multiple medications for diabetes treatment (21). Notably, this is a less restrictive definition of adherence, which we showed adherence estimation increases with adherence as more medications are prescribed. This approach effectively rewards the prescribing of increasing number of medications, creating increased opportunity for a patient to be regularly filling at least one of them. Conversely, the number of medications would have less impact if an average measure was used, while a more restrictive adherence measure would create a dis-incentive to increasing the number of medications prescribed.

We originally hypothesized that more restrictive PDC definitions for multiple medications would be more strongly associated with improved heart failure outcomes. In clinical trials, GDMT has been shown to improve hospitalization rates and mortality for patients with HFrEF and one study found an association between PDC measurement and improved HFrEFF, specifically all-cause hospitalizations (18,19). However, we found that higher PDC was associated with increased rates of ED visits, hospitalizations, and death regardless of which definition was used. There are several potential reasons for this unexpected association. First, our principal aim of the study was the measurement of adherence to multiple medications. Our cohort adherence measure was developed to assess this relationship; for instance, we allowed for up titration of GDMT before time zero to allow for a greater number of medications per patient. It is possible that the alteration of time zero may have introduced bias which possibly reflected sicker patients being more adherent to therapy as their chronic condition progresses (22). Second, we estimated PDC over a six-month period and then followed patients for outcomes over months, not years, which may not have allowed for sufficient time to observe the impact of adherence on clinical outcomes. Prior PDC outcome analyses have employed longer time periods for PDC calculation (i.e. one year v. six months) and, importantly, longer follow-up times (18,23,24). Third, the benefit of GDMT has mostly been shown in the HFrEF population. Notably, when performing our analysis for this subgroup we found no associated harm with increased adherence.

Limitations of our study, in addition to the ones explored above, include those inherent to all adherence measures: the lack of ability to directly measure adherence. This is an inherent flaw of measuring adherence at the population level. However, we calculated PDC with a technique that was similar to prior work and in the way advocated for by the PQA (9,11,18,25). Additionally, our outcome analysis was limited by its retrospective approach as this limited ability to record other patient-level factors that affect hospitalizations and outcomes of heart failure such as dietary habits, other illnesses, and exercise tendencies. Furthermore, this study took place in a single health system, which might limit generalizability. Lastly, we employed heart failure ICD codes for inclusion in our study which can be suboptimal for true characterization of disease (26).

### Conclusion

The four classes of GDMT for heart failure are known to improve patient health outcomes. Despite this, there is considerable variation in how to measure and assess adherence to a complex medication regimen at a population level. We described seven distinct methods of calculating adherence for GDMT and found considerable variation in estimates of adherence based on PDC. These differences suggests that adherence measures for multiple medications must be thoughtfully applied to the intervention, policy, or clinical context for which it is being utilized.

## Data Availability

Data will be available upon request.

